# Frequency and Medical Costs of Hypersensitivity- and Anaphylaxis-Related Adverse Events for Different Intravenous Iron Products Using the US Food and Drug Administration Adverse Event Reporting System (FAERS)

**DOI:** 10.64898/2026.04.30.26352160

**Authors:** Ye Wang, Syed Numan

## Abstract

**Background:** In the United States (US), several intravenous (IV) iron products are available for treatment of iron deficiency, including low-molecular-weight iron dextran (LMWID), iron sucrose (IS), ferumoxytol (FM), ferric carboxymaltose (FCM), ferric derisomaltose (FD), and sodium ferric gluconate (FG). However, these IV iron products are associated with rare, but serious, hypersensitivity and anaphylactic reactions.

**Objective:** This study aimed to assess the frequencies of hypersensitivity and anaphylactic reactions and associated downstream medical costs of the six IV iron products in the US.

**Methods:** This study used data from the US Food and Drug Administration (FDA) Adverse Event Reporting System (FAERS) from January 1, 2014, to June 30, 2024. The lower bound of the 90% confidence interval of the reporting odds ratio (ROR05) was used to identify a likely drug–adverse event (AE) association related to hypersensitivity and anaphylactic reactions. Downstream medical costs were estimated using Agency for Healthcare Research and Quality/Healthcare Cost and Utilization Project data.

**Results:** Signal strength of a likely drug-AE association for hypersensitivity was highest with FG (ROR05=9.66) and lowest with FCM (2.87). Signal strength for anaphylactic reactions was highest with FM (43.59) and lowest with FCM (6.99). The medical cost per AE was lowest with FCM (US$2,348) and highest with LMWID ($9,593).

**Conclusion:** FCM had the lowest signal strength of a likely drug–AE association for hypersensitivity and anaphylaxis and the lowest medical cost per AE in the US patient population, demonstrating its potential value by improving patient safety while lowering overall medical spending.

**Plain Language Summary:** This study has found that ferric carboxymaltose (FCM) had the lowest signal strength of a likely drug-adverse event (AE) association for hypersensitivity and anaphylaxis, compared to other intravenous iron products. FCM also had the lowest downstream medical cost per AE in the US patient population. These findings suggest that FCM may provide value by improving patient safety while reducing overall medical spending in the real-world setting.

## Introduction

Iron deficiency (ID) affects around 2 billion people globally^1^ and is the most common cause of anemia, accounting for approximately 66% of all cases worldwide.^2^ According to US National Health and Nutrition Examination Survey (NHANES), the estimated overall prevalence of ID among adults was 14%.^3^ ID is often associated with substantial negative sequelae, including fatigue, cognitive dysfunction, and reductions in work performance, with more severe symptoms likely in those with iron deficiency anemia (IDA).^4^

Management of IDA involves treatment of underlying causes (e.g., gastrointestinal bleeding) along with oral iron supplementation.^5,6^ However, oral iron therapy is associated with poor gastrointestinal tolerability and absorption, resulting in limited treatment response.^5,6^ Therefore, intravenous (IV) iron products may be required.^5,6^

Currently, there are six IV iron products in the US—low-molecular-weight iron dextran (LMWID), iron sucrose (IS), ferumoxytol (FM), ferric carboxymaltose (FCM), ferric derisomaltose (FD), and sodium ferric gluconate complex in sucrose (FG).^7^ Among them, FM, FCM and FD are used increasingly more often due to their increased stability and improved safety.^8-10^ However, there is limited evidence for different IV iron products with regard to the risk of hypersensitivity and anaphylactic reactions, largely due to the low incidence of these serious adverse events (AEs).^10-12^ Some real-world evidence suggests that there may be differences between available IV iron products, but this is inconsistent.^13,14^ Given the rarity of IV iron-related hypersensitivity and anaphylactic reactions, and insufficient evidence on the newest IV iron products, there is a need to assess these AEs among the IV iron products. Therefore, this study aims to evaluate the frequency and downstream medical costs of hypersensitivity and anaphylaxis related AEs for different IV iron products.

## Methods

### Data Source

The US Food and Drug Administration (FDA) Adverse Event Reporting System (FAERS) is a database of AEs submitted by healthcare professionals, consumers, and manufacturers that is designed to support post-marketing safety surveillance and may be used to identify potential safety concerns.^15^ Quarterly reports of FAERS data can be accessed from the FDA website, using RxFilter^®^ data optimization technology with established methods for de-duplication and mapping of drug names to standardized vocabularies.^16,17^

Post-marketing case reports submitted to the FAERS between January 1, 2014, and June 30, 2024, were extracted for six branded and generic IV iron products (FCM, FD, FG, FM, LMWID and IS) regarding hypersensitivity and anaphylactic reactions in the US patient population. These AEs were defined according to the Standardized Medical Queries (SMQs) in the Medical Dictionary for Regulatory Activities (MedDRA^®^), version 22.0. Data validation included verification that all case reports contained completed key identification fields, such as patient identifier, case identifier (individual patients could be associated with more than one case report), drug sequence, and AE terms in MedDRA.^16^ Case reports with incorrect or missing fields were excluded.^16^ Drug mapping and de-duplication of case reports were then completed.^16^

### Disproportionality Analysis

For each IV iron product, overall primary suspect case counts and proportion of cases leading to hospitalizations and deaths were reported. Primary suspect cases were defined as case reports where the IV iron product was designated as the most likely cause of the AEs of interest.

Reporting odds ratios (RORs) were used to determine the strength of the association between each IV iron product and the AEs of interest.^17^ The ROR is the pharmacovigilance measure that corresponds to the odds ratio and is less likely to lead to biased estimates compared to other disproportionality measures.^18^ It is defined as the odds of the AE of interest occurring in the case reports for the IV iron product of interest, divided by the odds of the AE of interest occurring in the case reports for patients receiving any other drug administered by any route. The odds of the AE of interest are the probability that the AE would occur divided by the probability that the AE would not occur. The 90% confidence interval (CI) limits of the ROR were established using an approximation of the normal distribution.^19^ The lower bound of the ROR distribution (ROR05) provided 95% certainty that the true mean of the population was at or above the reported number. A signal of a likely drug–AE association was indicated if ROR05 was ≥1 with at least 5 case counts. The masking ratio for the lower bound of the 90% confidence interval (MRCI) was calculated to determine whether there was any masking effect of disproportionate signals due to the presence of a disproportionate signal for another drug in the database with the same AE of interest.^20^ The ROR05 MRCI was equal to the ROR05 using the comparison group for the ROR without all other IV iron products divided by the ROR05 using the comparison group for the ROR with all other IV iron products. An ROR05 masking ratio that was substantially greater than 1.0 was considered indicative of masking.

A sensitivity analysis was performed using all suspect cases, defined as all case reports where the IV iron product was mentioned, including cases where it was a primary suspect, a secondary suspect, a concomitant medication, an interacting medication, or where the “suspect” field in FAERS was incomplete (i.e., not specified).

### Cost Analysis

Direct downstream medical costs associated with hypersensitivity and anaphylactic reactions were estimated for each IV iron product. The International Classification of Diseases, 10th Revision, Clinical Modification (ICD-10-CM) codes were used to identify diagnoses and procedures associated with the AEs of interest from the Agency for Healthcare Research and Quality/Healthcare Cost and Utilization Project (AHRQ/HCUP),^21-23^ which were then mapped to the preferred terms (PTs) within each SMQ category in MedDRA from the FAERS case reports. Only PTs that can be cross walked to ICD codes were included in the cost analysis. The downstream medical costs include cost associated with death, non-death hospitalization inpatient cost, and emergency room (ER) cost.^17,24^ The cost associated with death was estimated by multiplying the frequency of deaths related to mapped PTs and the average cost of a hospitalization that ended in death.^25^ The inpatient cost for each IV iron product was estimated by multiplying the frequency of AEs identified from mapped PTs that resulted in hospitalization with the average cost of AE-related hospitalization. The ER cost was calculated by multiplying the frequency of AEs that occurred in an ER with the average cost of AE-related ER visits. These costs were then added up to generate the total downstream medical cost for each IV iron product, which was further divided by the total number of AEs related to each IV iron product to obtain the average cost per AE. All costs are presented in US dollars and were adjusted for inflation on a yearly basis using the medical care services component of the consumer price index for December of each year.

## Results

In total, 9,482,774 FAERS case reports were available for the six IV iron products between January 1, 2014, and June 30, 2024, of which there were no cases with missing MedDRA AE terms. Therefore, the primary and total suspect case count analysis covered 100% of case reports for the overall ROR05 estimates. However, FAERS outcome field data (i.e., hospitalization and death) were missing for 5,775,009 cases (60.9%). Therefore, calculation of proportions and ROR05 values for associated hospitalizations and deaths was based on 39.1% coverage (3,707,765 cases). All masking ratio values were close to 1.0, indicating no evidence of masking (Supplementary Table 1).

### Hypersensitivity Reactions

In the primary analysis, all six IV iron products were found to have a likely drug–AE association for hypersensitivity (Table 1 and Figure 1). The signal strength was highest for FG (ROR05: 9.66; case count: 284), followed by IS (7.07; 613), FM (6.56; 387), LMWID (6.17; 177), and FD (4.62; 165), with FCM being the lowest (2.87; 727). All IV iron products had a likely drug–AE association for hypersensitivity resulting in hospitalization, with the highest signal strength for FG (14.50; 73) and the lowest for FCM (1.55; 54). For primary suspect cases of hypersensitivity resulting in death, a likely drug–AE association was found for LMWID (21.68; 13), FM (17.71; 21), FG (4.95; 5), and IS (2.38; 6). No signal was found for FCM (0.21; 2) or FD (0.44; 1).

**Table 1.** Primary Suspect Case Counts and Disproportionality for Hypersensitivity and Anaphylactic Reactions.

| IV iron | Hypersensitivity |  | Anaphylactic reaction |  |
| --- | --- | --- | --- | --- |
|  | Cases, n (%)<sup>a</sup> | ROR05 | Cases, n (%)<sup>a</sup> | ROR05 |
| FCM |  |  |  |  |
| Primary suspect cases | 727 (100.0) | 2.87 | 81 (100.0) | 6.99 |
| Hospitalizations | 54 (7.4) | 1.55 | 11 (13.6) | 1.86 |
| Deaths | 2 (0.3) | 0.21 | 1 (1.2) | 0.24 |
| FD |  |  |  |  |
| Primary suspect cases | 165 (100.0) | 4.62 | 22 (100.0) | 10.89 |
| Hospitalizations | 14 (8.5) | 2.18 | 5 (22.7) | 4.48 |
| Deaths | 1 (0.6) | 0.44 | 1 (4.6) | 1.59 |
| FG |  |  |  |  |
| Primary suspect cases | 284 (100.0) | 9.66 | 62 (100.0) | 34.09 |
| Hospitalizations | 73 (25.7) | 14.50 | 19 (30.7) | 22.49 |
| Deaths | 5 (1.8) | 4.95 | 3 (4.8) | 8.67 |
| FM |  |  |  |  |
| Primary suspect cases | 387 (100.0) | 6.56 | 126 (100.0) | 43.59 |
| Hospitalizations | 87 (22.5) | 9.76 | 46 (36.5) | 36.54 |
| Deaths | 21 (5.4) | 17.71 | 13 (10.3) | 35.89 |
| LMWID |  |  |  |  |
| Primary suspect cases | 177 (100.0) | 6.17 | 49 (100.0) | 32.78 |
| Hospitalizations | 41 (23.2) | 9.20 | 21 (42.9) | 32.07 |
| Deaths | 13 (7.3) | 21.68 | 6 (12.2) | 28.88 |
| IS |  |  |  |  |
| Primary suspect cases | 613 (100.0) | 7.07 | 76 (100.0) | 15.24 |
| Hospitalizations | 118 (19.3) | 8.79 | 27 (35.5) | 12.65 |
| Deaths | 6 (1.0) | 2.38 | 2 (2.6) | 1.75 |

<sup>a</sup>The percentages for hospitalizations and deaths were calculated based on the number of primary suspect cases for each IV iron product.
Abbreviations: *FCM*: ferric carboxymaltose; *FD*: ferric derisomaltose; *FG*: sodium ferric gluconate; *FM*: ferumoxytol; *IS*: iron sucrose; *IV*: intravenous; *LMWID*: low-molecular-weight iron dextran; *ROR05*: lower bound of the 90% confidence interval of reporting odds ratio.

**Figure 1.**
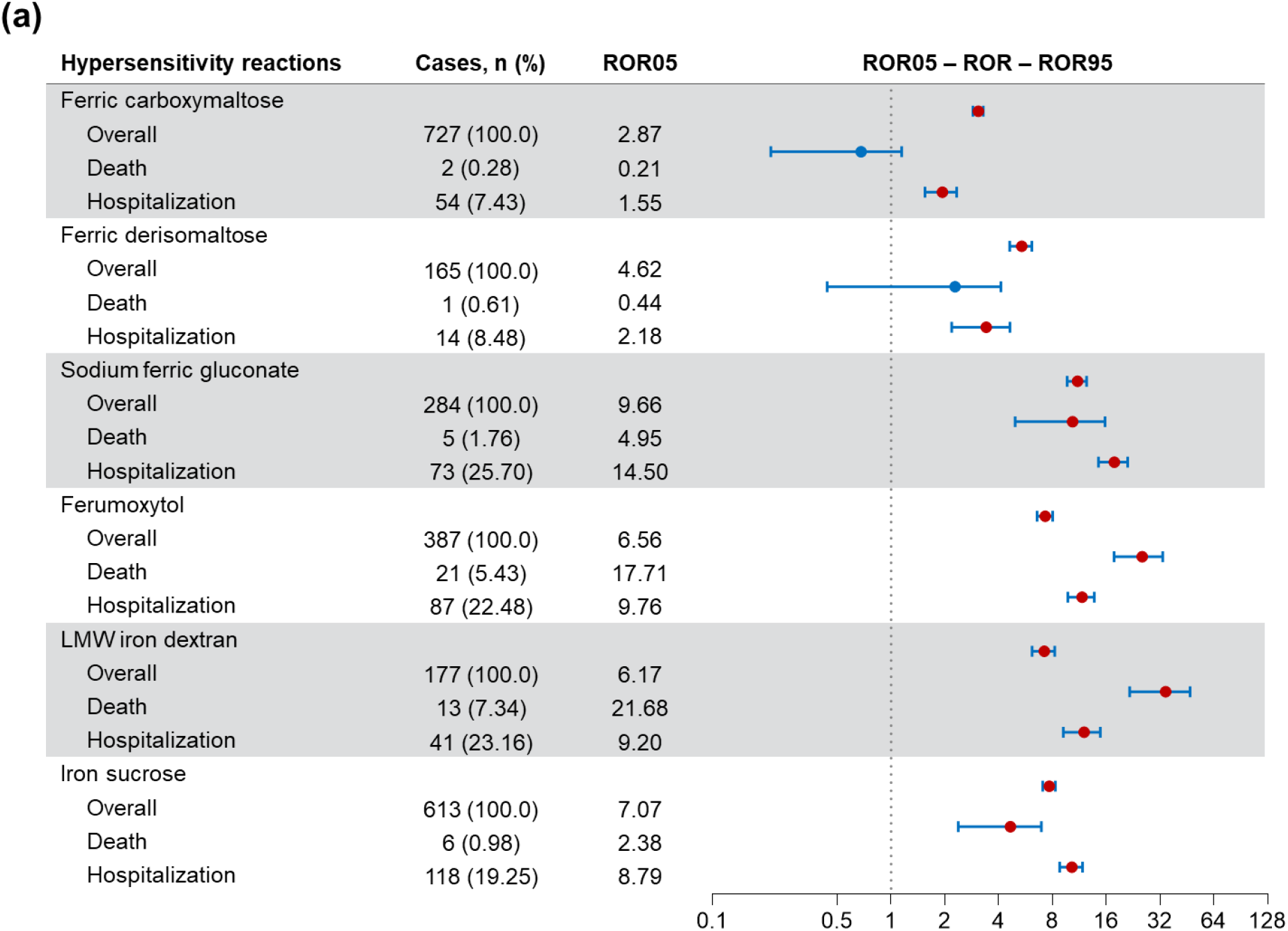
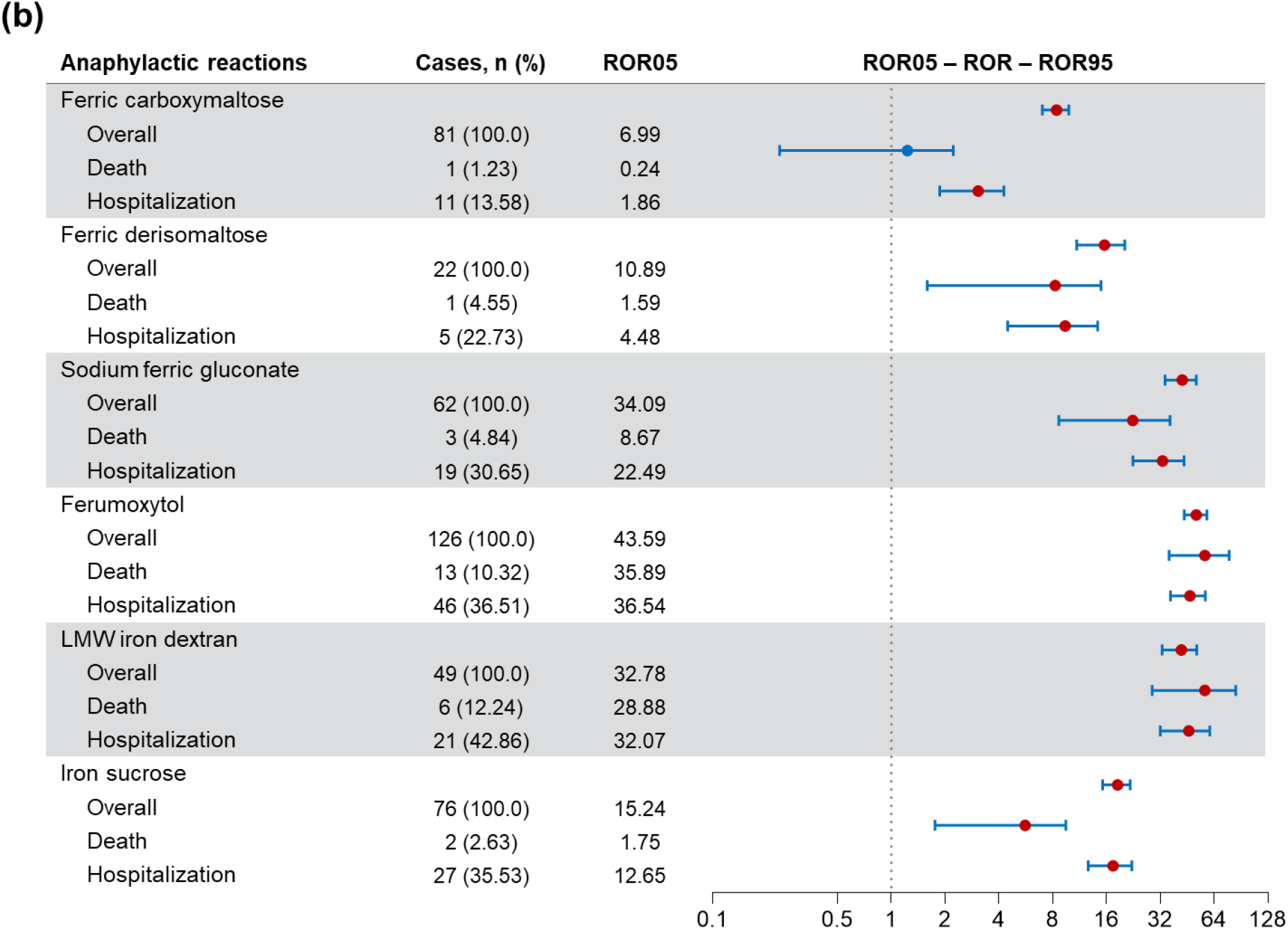
Disproportionality analysis^a^ for primary suspect cases with each of the six IV iron products for (a) hypersensitivity and (b) anaphylactic reactions. ^a^The error bars around ROR represent the upper (ROR95) and lower (ROR05) bounds of the 90% confidence interval. AEs with ≥5 cases and an ROR05 ≥1 indicated a signal for drug–AE association. *AE*: adverse event; *IV*: intravenous; *LMW*: low molecular weight; *ROR*: reporting odds ratio; *ROR05*: lower bound of the 90% confidence interval of reporting odds ratio; *ROR95*: upper bound of the 90% confidence interval of reporting odds ratio.

### Anaphylactic Reactions

All six IV iron products had a likely drug–AE association for anaphylactic reaction (Table 1, Figure 1). The signal strength was highest for FM (ROR05: 43.59; case count: 126), followed by FG (34.09; 62), LMWID (32.78; 49), IS (15.24; 76), and FD (10.89; 22). FCM had the lowest signal strength (6.99; 81). A likely drug–AE association for anaphylactic reaction resulting in hospitalization was also detected for all six IV iron products, with the signal strength being highest with FM (36.54; 46) and lowest with FCM (1.86; 11). For anaphylactic reaction resulting in death, only FM (35.89; 13) and LMWID (28.88; 6) showed a likely drug–AE association. While the ROR05 values were ≥1 for FD (1.59), FG (8.67), and IS (1.75), the number of deaths was <5 for each of these IV iron products (i.e., 1, 3, and 2 deaths, respectively). Therefore, the prespecified criteria for a likely drug–AE association were not met. No signal was found for FCM (0.24;1).

### Sensitivity Analysis

In the sensitivity analysis with all suspect cases, all six IV iron products had a likely drug–AE association for hypersensitivity (Supplementary Table 2). The signal strength was highest for IS (ROR05: 7.51; case count: 1312), followed by FG (7.42; 308), FM (5.51; 409), and FD (4.54; 165). FCM had the lowest signal strength (2.81; 754). A likely drug–AE association for hypersensitivity resulting in hospitalization was also observed with all six IV iron products, with FG having the highest signal strength (13.21; 84), followed by FM (8.88; 94), IS (6.94; 192), LMWID (6.83; 45), and FD (2.16; 14) and FCM (1.62; 59). For hypersensitivity resulting in death, a likely drug–AE association was found only for LMWID (17.54; 15), FM (15.73; 22), FG (7.31; 8), and IS (5.76; 22). No signal was found for FCM (0.37; 3) or FD (0.43; 1).

All six IV iron products displayed a likely drug–AE association for anaphylactic reaction (Supplementary Table 2). The highest signal strength was found for FM (ROR05: 36.32; case count: 127), followed by FG (27.35; 64), LMWID (22.20; 50), FD (10.77; 22), and IS (9.75; 101). FCM had the lowest signal strength (6.88; 84). For anaphylactic reactions leading to hospitalizations, the highest signal strength was observed for FM (31.34; 47), followed by LMWID (22.87; 22), FG (18.79; 20), IS (7.82; 34), FD (4.43; 5), and FCM (1.76; 11). In the case of anaphylactic reactions resulting in death, the highest signal strength was seen for FM (30.16; 13), followed by LMWID (19.64; 6). While the ROR05 values were ≥1 for FG (10.37), FD (1.57) and IS (1.55), the number of deaths was <5 for each of these IV iron products (i.e., 4, 1, and 3 deaths, respectively). FCM did not have a likely drug–AE association for anaphylactic reaction resulting in death (0.73; 2).

### Downstream Medical Costs

Downstream medical costs associated with managing hypersensitivity and anaphylactic reactions for the six IV iron products are shown in Table 2. FM had the highest total medical cost ($2,570,480) associated with hypersensitivity and anaphylactic reactions, while FD had the lowest total medical cost ($611,135).

**Table 2.**
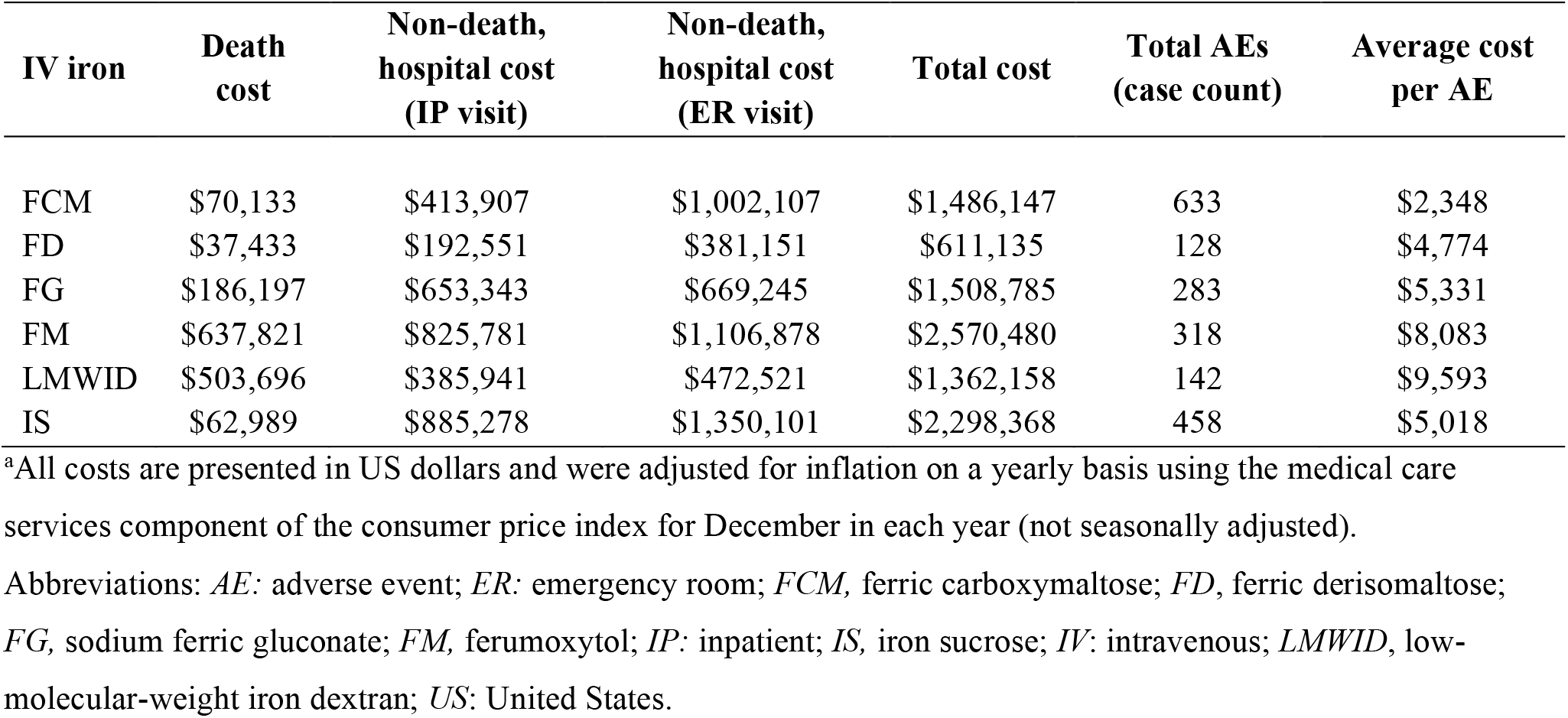
Downstream Medical Costs Associated with Managing Hypersensitivity and Anaphylactic Reactions with IV Iron Products ^a^.

| IV iron | Death cost | Non-death, hospital cost (IP visit) | Non-death, hospital cost (ER visit) | Total cost | Total AEs (case count) | Average cost per AE |
| --- | --- | --- | --- | --- | --- | --- |
| FCM | \$70,133 | \$413,907 | \$1,002,107 | \$1,486,147 | 633 | \$2,348 |
| FD | \$37,433 | \$192,551 | \$381,151 | \$611,135 | 128 | \$4,774 |
| FG | \$186,197 | \$653,343 | \$669,245 | \$1,508,785 | 283 | \$5,331 |
| FM | \$637,821 | \$825,781 | \$1,106,878 | \$2,570,480 | 318 | \$8,083 |
| LMWID | \$503,696 | \$385,941 | \$472,521 | \$1,362,158 | 142 | \$9,593 |
| IS | \$62,989 | \$885,278 | \$1,350,101 | \$2,298,368 | 458 | \$5,018 |

<sup>a</sup>All costs are presented in US dollars and were adjusted for inflation on a yearly basis using the medical care services component of the consumer price index for December in each year (not seasonally adjusted).
Abbreviations: *AE*: adverse event; *ER*: emergency room; *FCM*, ferric carboxymaltose; *FD*, ferric derisomaltose; *FG*, sodium ferric gluconate; *FM*, ferumoxytol; *IP*: inpatient; *IS*, iron sucrose; *IV*: intravenous; *LMWID*, low-molecular-weight iron dextran; *US*: United States.

In the cost per AE analysis, FCM was associated with the lowest average cost per AE (US$2,348). The highest cost per AE was observed with LMWID (US$9,593).

## Discussion

This study evaluated the frequency and downstream medical cost related to hypersensitivity and anaphylactic reactions for the six IV iron products in the US patient population using real-world data. FCM was found to have the lowest overall signal strength for a likely drug–AE association related to hypersensitivity and anaphylactic reaction and the lowest proportions of associated hospitalizations and deaths. Although clinical trials are important for evaluation of drug efficacy and safety, they often have strict selection criteria, leading to a homogeneous cohort of patients. They also tend to exclude patients more likely to experience AEs, leading to small sample sizes insufficient to detect rare yet important AEs. In contrast, studies utilizing real-world data allow us to investigate the safety profiles of IV iron products in broader, more heterogeneous patient populations in real-world settings.

This study has found that all six IV iron products were associated with risks of hypersensitivity and anaphylactic reactions; however, differences in signal strength among products and in the proportionality for these AEs associated with hospitalization and death were observed. Specifically, the drug–AE association signals for both hypersensitivity and anaphylactic reactions were lowest with FCM. Similarly, the signals were also the lowest with FCM for hypersensitivity and anaphylactic cases leading to hospitalization and no signal was observed for cases leading to death. In contrast, FG had the highest drug–AE association signal for hypersensitivity and associated hospitalizations, while the highest signal for hypersensitivity-associated death was observed with LMWID. For anaphylactic reactions, FM had the highest drug–AE association signal for overall primary cases, and anaphylactic reaction-associated hospitalizations and death. Sensitivity analyses using total suspect cases have generated consistent findings, with FCM having the lowest drug–AE association across all metrics. The findings of this study are consistent with those from a previous study that used data from FAERS to assess four IV iron products (i.e., LMWID, IS, FM and FCM) between January 1, 2014, and December 31, 2019.^17^ It was found that FCM had the lowest signal strength for overall hypersensitivity related AEs and associated hospitalizations. FCM also had the lowest signal strength for overall anaphylaxis related AEs and associated hospitalizations. FCM showed no signal for a likely association with hypersensitivity or anaphylaxis resulting in death.

The downstream medical costs associated with hypersensitivity and anaphylactic reactions with IV iron products were also analyzed in the real-world context to identify the products associated with the greatest economic benefits. These estimates provide an alternative way to assess safety risks posed by drugs based on AEs and associated costs, and act as an accessible real-world reference for post-marketing drug safety.^24^ Because drugs approved earlier tend to accumulate greater downstream medical costs, the average cost per AE was calculated to account for differences in time on the market for different drugs. It has been found that LMWID had the highest average cost per AE. In contrast, FCM had the lowest average cost per AE, indicating that FCM had the lowest economic burden associated with hypersensitivity and anaphylactic reactions.

## Limitations

This study has a few limitations. First, this study utilized data solely from the US patient population. Therefore, the findings of this study are exclusive to patients in the US and are not generalizable to patient populations from other countries. Second, although the FAERS suspect case analyses had full data coverage, there was a high proportion of missing data for analyses of associated hospitalizations and deaths. How the missing data affects the findings related to hospitalizations and deaths is unknown. Third, the FAERS database provides event-level rather than patient-level information; therefore, only event-level reporting rates (i.e., event counts) can be estimated. Patient counts and patient characteristics cannot be generated. In addition, AE reporting rates may be influenced by time-on-market, i.e., more recently approved drugs may have lower reporting rates. Comparison across drugs needs to be interpreted with caution. Finally, the designation of primary suspect cases in the FAERS database is subjective. However, this limitation was mitigated in the current study by the sensitivity analysis of the total suspect case reports, which showed similar findings to the primary analysis.

## Conclusion

FCM showed the lowest signal strength of a likely drug–AE association related to hypersensitivity and anaphylactic reactions, and the lowest drug–AE association signals for related hospitalizations and deaths, among the six IV iron products. FCM also had the lowest downstream medical costs per AE. The findings suggest that FCM may be associated with a reduced healthcare and cost burden among individuals who required IV iron products, demonstrating its potential value by improving patient safety while lowering overall medical spending.

## Data Availability

All data produced in the present study are available upon reasonable request to the authors.

---

**Supplementary Table 1.**
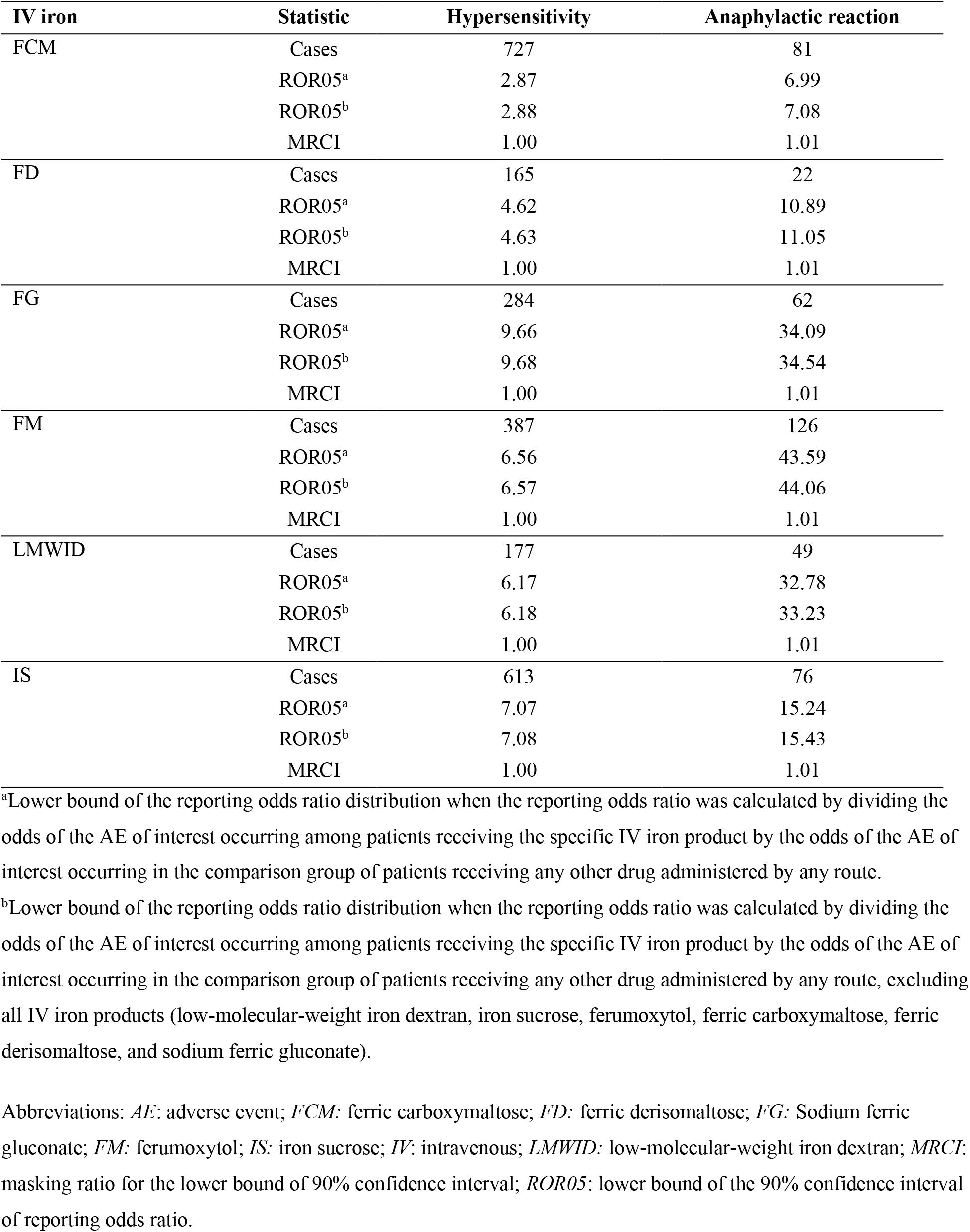
Calculation of ROR05 Masking Ratios Derived from Primary Suspect Cases of Hypersensitivity or Anaphylactic Reaction Associated with IV Iron Products.

| IV iron | Statistic | Hypersensitivity | Anaphylactic reaction |
| --- | --- | --- | --- |
| FCM | Cases | 727 | 81 |
|  | ROR05<sup>a</sup> | 2.87 | 6.99 |
|  | ROR05<sup>b</sup> | 2.88 | 7.08 |
|  | MRCI | 1.00 | 1.01 |
| FD | Cases | 165 | 22 |
|  | ROR05<sup>a</sup> | 4.62 | 10.89 |
|  | ROR05<sup>b</sup> | 4.63 | 11.05 |
|  | MRCI | 1.00 | 1.01 |
| FG | Cases | 284 | 62 |
|  | ROR05<sup>a</sup> | 9.66 | 34.09 |
|  | ROR05<sup>b</sup> | 9.68 | 34.54 |
|  | MRCI | 1.00 | 1.01 |
| FM | Cases | 387 | 126 |
|  | ROR05<sup>a</sup> | 6.56 | 43.59 |
|  | ROR05<sup>b</sup> | 6.57 | 44.06 |
|  | MRCI | 1.00 | 1.01 |
| LMWID | Cases | 177 | 49 |
|  | ROR05<sup>a</sup> | 6.17 | 32.78 |
|  | ROR05<sup>b</sup> | 6.18 | 33.23 |
|  | MRCI | 1.00 | 1.01 |
| IS | Cases | 613 | 76 |
|  | ROR05<sup>a</sup> | 7.07 | 15.24 |
|  | ROR05<sup>b</sup> | 7.08 | 15.43 |
|  | MRCI | 1.00 | 1.01 |

<sup>a</sup>Lower bound of the reporting odds ratio distribution when the reporting odds ratio was calculated by dividing the odds of the AE of interest occurring among patients receiving the specific IV iron product by the odds of the AE of interest occurring in the comparison group of patients receiving any other drug administered by any route.
<sup>b</sup>Lower bound of the reporting odds ratio distribution when the reporting odds ratio was calculated by dividing the odds of the AE of interest occurring among patients receiving the specific IV iron product by the odds of the AE of interest occurring in the comparison group of patients receiving any other drug administered by any route, excluding all IV iron products (low-molecular-weight iron dextran, iron sucrose, ferumoxytol, ferric carboxymaltose, ferric derisomaltose, and sodium ferric gluconate).
Abbreviations: *AE*: adverse event; *FCM*: ferric carboxymaltose; *FD*: ferric derisomaltose; *FG*: Sodium ferric gluconate; *FM*: ferumoxytol; *IS*: iron sucrose; *IV*: intravenous; *LMWID*: low-molecular-weight iron dextran; *MRCI*: masking ratio for the lower bound of 90% confidence interval; *ROR05*: lower bound of the 90% confidence interval of reporting odds ratio.

**Supplementary Table 2.** Sensitivity Analysis of All Suspect Case Counts and Disproportionality Associated with Hypersensitivity and Anaphylactic Reactions.

| IV iron | Hypersensitivity |  | Anaphylactic reaction |  |
| --- | --- | --- | --- | --- |
|  | Cases, n (%)<sup>a</sup> | ROR05 | Cases, n (%)<sup>a</sup> | ROR05 |
| FCM |  |  |  |  |
| All suspect cases | 754 (100.0) | 2.81 | 84 (100.0) | 6.88 |
| Hospitalizations | 59 (7.8) | 1.62 | 11 (13.1) | 1.76 |
| Deaths | 3 (0.4) | 0.37 | 2 (2.4) | 0.73 |
| FD |  |  |  |  |
| All suspect cases | 165 (100.0) | 4.54 | 22 (100.0) | 10.77 |
| Hospitalizations | 14 (8.5) | 2.16 | 5 (22.7) | 4.43 |
| Deaths | 1 (0.6) | 0.43 | 1 (4.5) | 1.57 |
| FG |  |  |  |  |
| All suspect cases | 308 (100.0) | 7.42 | 64 (100.0) | 27.35 |
| Hospitalizations | 84 (27.3) | 13.21 | 20 (31.3) | 18.79 |
| Deaths | 8 (2.6) | 7.31 | 4 (6.3) | 10.37 |
| FM |  |  |  |  |
| All suspect cases | 409 (100.0) | 5.51 | 127 (100.0) | 36.32 |
| Hospitalizations | 94 (23.0) | 8.88 | 47 (37.0) | 31.34 |
| Deaths | 22 (5.4) | 15.73 | 13 (10.2) | 30.16 |
| LMWID |  |  |  |  |
| All suspect cases | 200 (100.0) | 4.27 | 50 (100.0) | 22.20 |
| Hospitalizations | 45 (22.5) | 6.83 | 22 (44.0) | 22.87 |
| Deaths | 15 (7.5) | 17.54 | 6 (12.0) | 19.64 |
| IS |  |  |  |  |
| All suspect cases | 1312 (100.0) | 7.51 | 101 (100.0) | 9.75 |
| Hospitalizations | 192 (14.6) | 6.94 | 34 (33.7) | 7.82 |
| Deaths | 22 (1.7) | 5.76 | 3 (3.0) | 1.55 |

<sup>a</sup>The percentages for hospitalizations and deaths were calculated based on the number of total suspect cases for each IV iron product.
Abbreviations: *FCM*: ferric carboxymaltose; *FD*: ferric derisomaltose; *FG*: Sodium ferric gluconate; *FM*: ferumoxytol; *IS*: iron sucrose; *IV*: intravenous; *LMWID*: low-molecular-weight iron dextran; *ROR05*: lower bound of the 90% confidence interval of reporting odds ratio.

